# Erosive potential of energy drink modified by calcium formulations on dental enamel: an *in vitro* study

**DOI:** 10.1101/2025.06.18.25329903

**Authors:** Erik Vinícius Martins Jácome, Marquiony Marques dos Santos, Dayanne Monielle Duarte Moura, Patricia Bittencourt Santos, Maria do Socorro da Costa Inácio, Ana Clara Soares Paiva Torres

## Abstract

**Objectives:** To evaluate the erosive potential of an energy drink modified with different calcium formulations and concentrations on dental enamel.

**Methods:** This *in vitro* study used sixty dental enamel specimens, divided into 12 groups: traditional Red Bull^®^ energy drink modified with three concentrations each of calcium/phosphorus/potassium complex (0.71, 1.43, and 2.15g), dicalcium malate (0.83, 1.66, and 2.50g), and calcium citrate malate (1.26, 2.53, and 3.80g); unmodified traditional Red Bull^®^ energy drink; deionized water; and commercially available calcium-supplemented Life Mix Baixa Caloria^®^ orange juice. The pH of all drinks was measured using a pHmeter. Specimens were exposed to the drinks for two minutes. Surface roughness and microhardness were assessed before and after exposure using a rugosimeter and a Vickers microhardness tester, respectively.

**Results:** The addition of calcium/phosphorus/potassium complex increased the energy drink’s pH proportionally to the concentration added. Specimens exposed to modified drinks showed reduced roughness (p < 0.004) compared to control drinks. Drinks modified with 2.50g of dicalcium malate and 2.15g of the calcium/phosphorus/potassium complex significantly preserved enamel microhardness (p < 0.004) compared to the unmodified energy drink. Across all modifications, enamel surface microhardness loss was lower than in control groups, except for deionized water.

**Conclusion:** The addition of calcium formulations to the energy drink effectively reduced its erosive potential at all concentrations, with the calcium/phosphorus/potassium complex demonstrating the greatest protective effect.

## 1. Introduction

Dental erosion is a type of non-carious lesion characterized by the chemical dissolution of enamel and dentin, leading to the progressive destruction of tooth structure [1]. Acid exposure reduces the mechanical resistance and surface hardness of these tissues [2], initially causing partial demineralization and softening of enamel or dentin, and later progressing to erosive tooth wear due to mechanical forces such as toothbrushing [3–5].

Among the etiological factors of dental erosion, exposure to extrinsic acids is the most common [6]. The erosive impact of acidic foods and drinks is influenced by frequency and form of consumption, as well as by individual physiological factors including saliva composition, dental anatomy, and occlusion [6]. Energy drinks are especially harmful due to their high concentration of acidulants—substances that regulate pH, extend shelf life, and enhance product stability—contributing to their high acidity [7–10]. Other factors such as pH, titratable acidity, calcium and phosphate content, and temperature also influence their erosive potential and the resulting risk to oral health [6].

The consequences of dental erosion include the loss of enamel smoothness and brightness, formation of concavities and rounded dentin surfaces, thinning of incisal edges, reduced occlusal vertical dimension, and dentin exposure [11]. These changes can lead to dental sensitivity, aesthetic dissatisfaction, and in advanced cases, pulp involvement when tooth structure loss is extensive [12,13].

Incorporating calcium formulations into acidic drinks has been proposed as a viable, safe, and effective strategy for preventing and controlling dental erosion [14]. However, few studies have specifically evaluated the addition of calcium to energy drinks while considering formulation characteristics that could enhance their anti-erosive effect. Thus, further research is necessary to identify effective modifications, as improvements in the composition of these drinks could help preserve the oral health of consumers, particularly those with pre-existing dental wear who are more susceptible to the progression of erosive lesions.

Therefore, this study aimed to evaluate the erosive potential of an energy drink modified with different calcium formulations and concentrations on dental enamel. The null hypothesis was that modifying energy drinks with varying calcium concentrations would not alter their erosive potential compared to the control groups.

## 2. Materials and methods

### 2.1. Study design

This *in vitro* experimental study was conducted in accordance with the ethical principles outlined in the Declaration of Helsinki. The study was approved for execution by the Research Ethics Committee of the Universidade do Estado do Rio Grande do Norte on May 15, 2023, under protocol number: 6.059.387.

### 2.2. Preparation of enamel specimens

To determine the minimum number of enamel specimens required for the experimental tests, a sample calculation was performed using G*Power software (version 3.1). The effect sizes for surface roughness and microhardness, calculated based on a model study, were 2.36 and 2.20, respectively. The calculation also considered an alpha error of 0.05% and a beta power of 95%, resulting in a requirement of five specimens per group, totaling 60 specimens.

The teeth used in this study were voluntarily donated by patients treated at the Dental Specialties Center in the municipality of Caraúbas, RN, Brazil. The attending dentist informed the principal investigator in advance of the scheduled days for molar extractions, which had been previously indicated by professionals from Primary Health Care Units. On these days, the researcher was present at the center and provided individual explanations about the study and the Informed Consent Form (ICF) to each patient in a private setting. The ICF was read in detail, with sufficient time allowed to answer questions, clarify the study objectives and potential risks, and ensure a free and informed decision. If needed, the participant was given additional time to decide. After agreeing to participate, patients signed the ICF and the Tooth Donation Form, authorizing the use of the extracted tooth/teeth for research purposes.

None of the donors belonged to a vulnerable population. Inclusion criteria were: individuals aged 18 years or older, receiving care at the Dental Specialties Center, and having a prior clinical indication for the extraction of one or more sound molars due to orthodontic, prosthetic, or surgical reasons (e.g., tooth impaction). Individuals with any mental condition impairing understanding of the study and informed decision-making were excluded. Therefore, we declare that none of the donors belonged to a vulnerable population and all donors provided written informed consent, which was given freely.

Between June 26 and August 4, 2023, twenty-six clinically healthy molars were collected from donations by twenty-three patients. The consent procedure was conducted exclusively by the principal investigator without the presence of other professionals, ensuring privacy and the absence of coercion. Donors were not compensated in any form, as the teeth were already scheduled for extraction due to independent clinical reasons and would otherwise have been discarded as biological waste according to the center’s biosafety protocols.

Each participant donated a maximum of two teeth. The use of human teeth instead of animal teeth in this study is scientifically and ethically justified. Scientifically, human teeth provide a substrate that is more representative of real clinical conditions, enhancing the external validity and applicability of the results to dental practice. Ethically, no institutionalized human tooth biobanks were available at the participating institutions. Therefore, the voluntary donation of teeth extracted for pre-existing clinical reasons represents a legal, acceptable, and respectful method for obtaining biological material, fully upholding donor autonomy.

Following extraction, the teeth were cleaned and stored in a 0.1% thymol solution at 4 °C until use. Enamel speimens were prepared from the flat, groove-free surfaces of the dental crowns. Each specimen measured 4 mm (width/length/thickness). To obtain the specimens, the crowns were sectioned using a double-sided diamond disc (American Burrs^®^, Palhoça, SC, Brazil) and a carborundum disc (American Burrs^®^, Palhoça, SC, Brazil), both mounted on a straight handpiece (Golgran^®^, São Paulo, SP, Brazil).

The enamel blocks were then flattened using a metallographic polisher (PL 02 E, Teclago, Vargem Grande Paulista, SP, Brazil) operating at high speed. Abrasive water sandpaper with grit sizes of 600, 800 (W-Max^®^, Betim, MG, Brazil) and 1,200 (Norton^®^, Black Ice, Guarulhos, SP, Brazil) was sequentially applied under deionized water cooling. Subsequently, the blocks were polished with a felt disc (Skill-Tec^®^, São Paulo, SP, Brazil), mounted on the polisher at high speed, without cooling, using a 0.5 μm diamond paste (Eagle Diamond, American Burrs^®^, Palhoça, SC, Brazil). To remove polishing residues, the specimens were immersed in deionized water and stirred for 10 minutes using a magnetic stirrer (752 A, Fisatom^®^, São Paulo, SP, Brazil).

Specimen quality was assessed using a microscope (Nikon, Eclipse MA100N, Tokyo, Japan) at 10X and 25X magnifications to ensure homogeneity. Specimens exhibiting alterations in color, texture, or enamel integrity were excluded. The final specimens presented a glassy, white appearance with no irregularities, scratches, or cracks in the central region designated for surface analysis.

### 2.3. Tested groups

The traditional energy drink Red Bull^®^ (Red Bull GmbH^®^, Manaus, AM, Brazil) was selected as the base for testing. This drink was modified with different concentrations and formulations of calcium, resulting in nine experimental formulations. The original (unmodified) energy drink was also tested as part of the negative control group, without calcium formulations. The negative control group also consisted of deionized water, which has a neutral pH and was obtained through ion removal (cations/anions) using an ion exchange resin system. For the positive control group, a commercially available calcium-supplemented orange juice (Life Mix Baixa Caloria^®^, São Paulo, SP, Brazil) was selected. This juice has a pH similar to an energy drink, with a difference in the calcium present in its composition. Table 1 shows the identification of the tested groups.

**Table 1.** Identification of tested groups.

| Group | Drink | Test substance added to drink | Concentration of test substance per ml of drink |
| --- | --- | --- | --- |
| EDCaPP0.71 | Red Bull® Energy Drink | Calcium/Phosphorus/Potassium Complex | 0,71 g / 50 ml |
| EDCaPP1.43 | Red Bull® Energy Drink | Calcium/Phosphorus/Potassium Complex | 1,43 g / 50 ml |
| EDCaPP2.15 | Red Bull® Energy Drink | Calcium/Phosphorus/Potassium Complex | 2,15 g / 50 ml |
| EDCaM0.83 | Red Bull® Energy Drink | Dicalcium Malate | 0,83 g / 50 ml |
| EDCaM1.66 | Red Bull® Energy Drink | Dicalcium Malate | 1,66 g / 50 ml |
| EDCaM2.50 | Red Bull® Energy Drink | Dicalcium Malate | 2,50 g / 50 ml |
| EDCaCM1.26 | Red Bull® Energy Drink | Calcium Citrate Malate | 1,26 g / 50 ml |
| EDCaCM2.53 | Red Bull® Energy Drink | Calcium Citrate Malate | 2,53 g / 50 ml |
| EDCaCM3.80 | Red Bull® Energy Drink | Calcium Citrate Malate | 3,80 g / 50 ml |
| ED | Red Bull® Energy Drink | -- | -- |
| DW | Deionized Water | -- | -- |
| OJCa | Orange juice with calcium Life Mix Baixa Caloria® | -- | -- |

The tested calcium compounds, added in the form of soluble powder, included: calcium/phosphorus/potassium complex at concentrations of 0.71g, 1.43g and 2.15g; dicalcium malate at concentrations of 0.83g, 1.66g and 2.50g; and calcium citrate malate at concentrations of 1.26g, 2.53g and 3.80g. These concentrations were established through mathematical calculations based on the raw calcium content in 500g of each compound and the required calcium levels in each tested concentration. The final calcium content was set at 300 mg, 600 mg, and 900 mg, respectively.

The selected calcium concentrations align with the limits established by the Brazilian Health Regulatory Agency, under RDC No. 719/2022 [15], which allows a maximum calcium addition of 1,000 milligrams in energy drinks. In addition, these concentrations fall within the safe daily calcium intake range, which suggests a maximum daily intake of 2,500 milligrams for individuals aged 19 to 50 [16].

To modify the energy drink, the calcium compounds were precisely weighed using an analytical balance (SHIMADZU^®^ TX323L; Kyoto, Japan) with a precision of 0.001 grams. Subsequently, 50mL of the energy drink was dispensed into 80 mL plastic containers (Mylabor^®^, São Paulo, SP, Brazil). The respective calcium compounds were added at the determined concentrations. The mixtures were manually homogenized for approximately three minutes using a glass stirring rod (Mylabor^®^, São Paulo, SP, Brazil) until the powder was fully dissolved.

### 2.4. Measuring of pH in drinks

Following preparations, the pH of each drink was measured using 50 mL of the sample at room temperature. A pHmeter (Quimis^®^ Q400AS; São Paulo, SP, Brazil) with a calibrated pH electrode was used for this analysis.

### 2.5. Exposure of enamel specimens to drinks

A preliminary pilot test was conducted using five additional enamel specimens to determine the optimal exposure time for demineralization. The specimens were progressively demineralized by immersion in an energy drink at room temperature. Surface microhardness was assessed after one-minute intervals of immersion. The results indicated that maximum enamel softening occurred after two minutes of immersion, as evidenced by a decrease in microhardness values. After three minutes of immersion, a slight increase in microhardness values was observed, suggesting partial loss of softened enamel structure. Based on these findings, an exposure time of two minutes was established for the experimental protocol.

To ensure a homogeneous distribution of specimens among the twelve experimental groups, the specimens were selected based on their initial surface microhardness values. A stratified random allocation was performed to evenly distribute specimens with lower and higher microhardness values across all groups. The specimens were ranked in ascending order of microhardness, and a random selection was conducted using the “RANDBETWEEN” function in Microsoft Excel (Microsoft Corporation^®^, Washington, United States).

Once the exposure time was defined and the specimens were randomized, 50 mL of each drink were dispensed into twelve separate 80 mL plastic containers. Five enamel specimens were placed in each container, ensuring complete immersion for two minutes, with their enamel surfaces facing upwards. At the end of this exposure period, the specimens were removed, immediately rinsed with deionized water to eliminate residual drinks components, and then air-dried for three minutes before proceeding with surface analyses.

### 2.6. Surface analysis of enamel specimens

All enamel specimens underwent surface roughness and microhardness analyses before and after exposure to the drinks. Enamel surface roughness was assessed using a rugosimeter (Mitutoyo^®^ Surftest SJ-210, Suzano, SP, Brazil). The device was calibrated according to the ISO 1997 reference standard [17] with the following parameters: sampling length (Cutoff) of 0.8mm; evaluation length of two times (x2); reading speed of 0.5 mm/s, and Ra as roughness parameter. Ten roughness measurements were performed per specimen (five before and five after exposure), ensuring a minimum spacing of 0.2 mm between each measurement and the specimen margins.

Superficial enamel microhardness was evaluated using a microhardness tester (Insize^®^ ISH-TDV2000-B, Boituva, SP, Brazil) with a Vickers pyramidal diamond indenter. Each specimen was positioned on the equipment table and adjusted using the eyepiece to ensure proper alignment. Ten indentations per specimen were made (five before and five after exposure) in the central enamel surface, with a minimum distance of 100 micrometers between indentations and 1000 micrometers from the edges. A static load of 100 g was applied for 15 seconds in the indentation on the enamel [18]. The percentage of surface microhardness loss (%SMHL) was calculated by averaging the initial and final microhardness values for each drink group. A flowchart of the experimental procedures is shown in Fig. 1.

**Fig 1.**
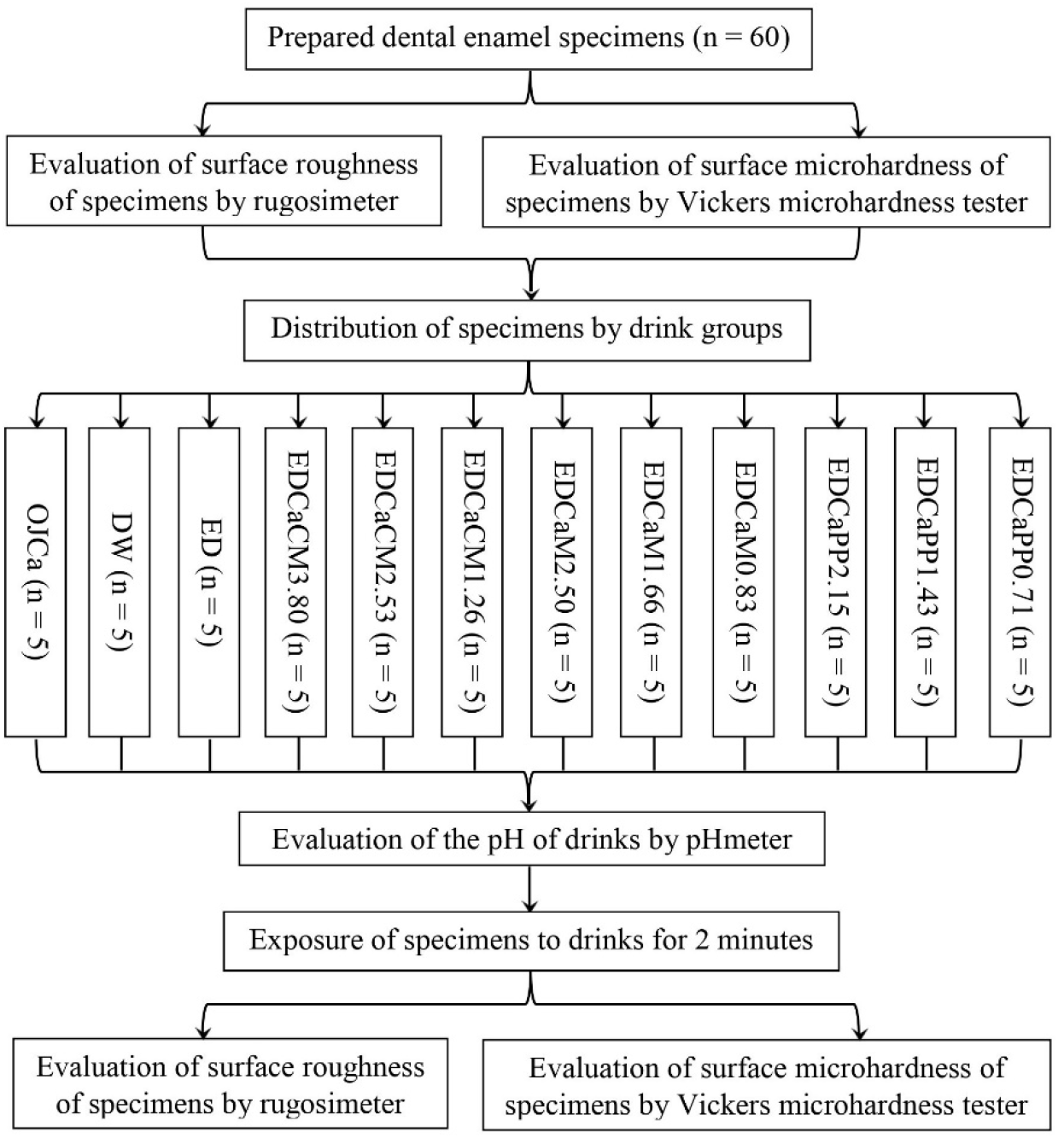
Flowchart of the experimental procedures. EDCaPP0.71: Red Bull^®^ Energy Drink + 0.71g of Calcium/Phosphorus/Potassium Complex; EDCaPP1.43: Red Bull^®^ Energy Drink + 1.43g of Calcium/Phosphorus/Potassium Complex; EDCaPP2.15: Red Bull^®^ Energy Drink + 2.15g of Calcium/Phosphorus/Potassium Complex; EDCaM0.83: Red Bull^®^ Energy Drink + 0.83g of Dicalcium Malate; EDCaM1.66: Red Bull^®^ Energy Drink + 1.66g of Dicalcium Malate; EDCaM2.50: Red Bull^®^ Energy Drink + 2.50g of Dicalcium Malate; EDCaCM1.26: Red Bull^®^ Energy Drink + 1.26g of Calcium Citrate Malate; EDCaCM2.53: Red Bull^®^ Energy Drink + 2.53g of Calcium Citrate Malate; EDCaCM3.80: Red Bull^®^ Energy Drink + 3.80g of Calcium Citrate Malate; ED: Red Bull^®^ Energy Drink; DW: Deionized Water; OJCa: Orange juice with calcium Life Mix Baixa Caloria^®^.

### 2.7. Data analysis

The data were analyzed quantitatively using Statistix software (Statistix version 8.0, Analytical Software, Florida, United States) with a significance level set at 5%. Initially, data normality was assessed using the Shapiro-Wilk test, which indicated a non-normal distribution for the roughness variable and a normal distribution for the microhardness variable. The mean was considered in the statistical tests for both variables, while the pH behavior of the drinks was analyzed descriptively.

For roughness analysis, the Wilcoxon test was applied to compare values before and after specimens exposure to the drinks. Subsequently, statistical differences between the drinks were assessed using the Kruskal-Wallis test, and when significance was detected, the Mann-Whitney test was used for pairwise comparisons, applying Bonferroni corrections with a p-value threshold of ≤ 0.004. For microhardness analysis, an Analysis of Variance (ANOVA) was performed to determine significant differences between time points (initial and final), the different drink groups, and their interaction. To identify which means were significantly different, the Tukey HSD (Honest Significant Difference) test was applied.

## 3. Results

The results were divided according to the variables considered in the study, the pH of the drinks, and the surface roughness and microhardness of the dental enamel.

### 3.1. pH of drinks

The pH analysis revealed that increasing the concentration of the calcium/phosphorus/potassium complex led to a rise in the pH of the energy drink. In contrast, both dicalcium malate and calcium citrate malate formulations showed an inverse trend, where increasing concentrations of these substances resulted in a decrease in the pH of the energy drink. Only the calcium/phosphorus/potassium complex concentrations raised the pH when compared to the unmodified energy drink (Fig. 2).

**Fig 2.**
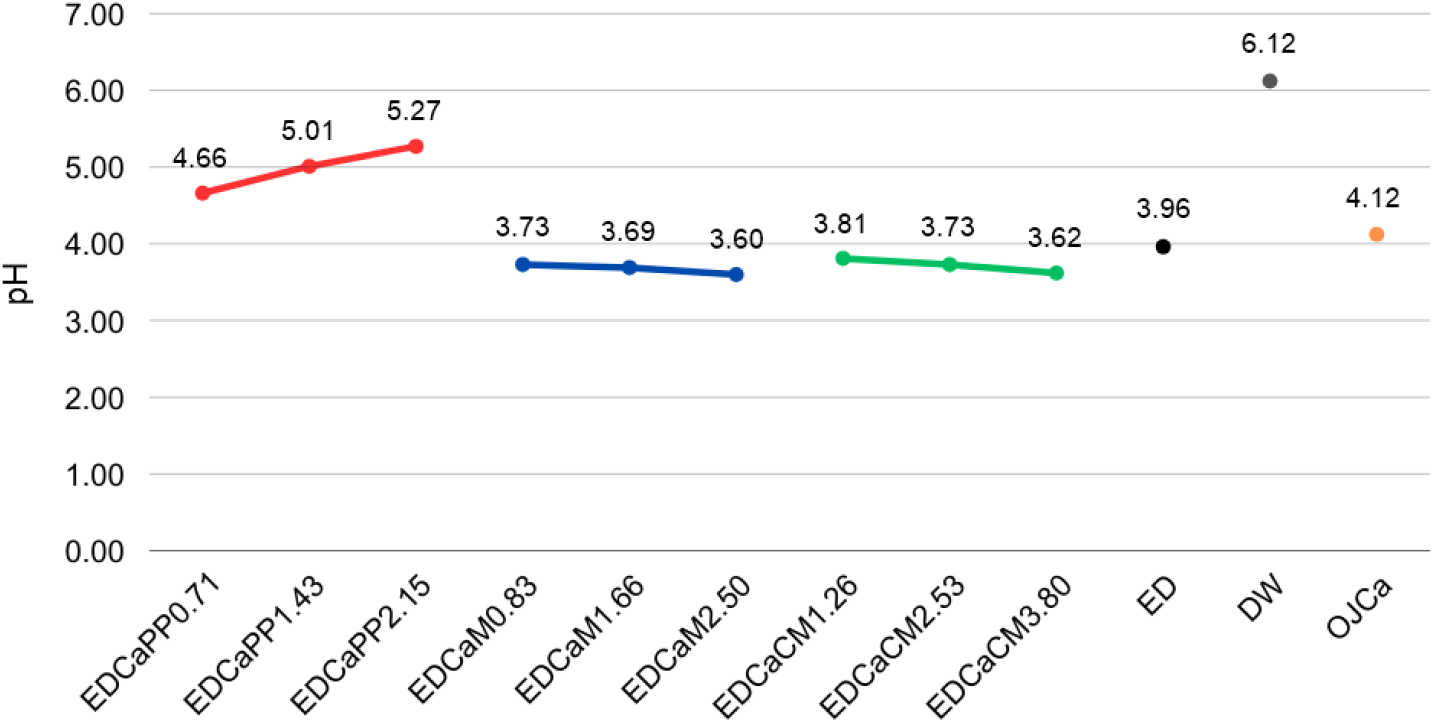
Line graph showing the pH behavior of the tested drinks. EDCaPP0.71: Red Bull^®^ Energy Drink + 0.71g of Calcium/Phosphorus/Potassium Complex; EDCaPP1.43: Red Bull^®^ Energy Drink + 1.43g of Calcium/Phosphorus/Potassium Complex; EDCaPP2.15: Red Bull^®^ Energy Drink + 2.15g of Calcium/Phosphorus/Potassium Complex; EDCaM0.83: Red Bull^®^ Energy Drink + 0.83g of Dicalcium Malate; EDCaM1.66: Red Bull^®^ Energy Drink + 1.66g of Dicalcium Malate; EDCaM2.50: Red Bull^®^ Energy Drink + 2.50g of Dicalcium Malate; EDCaCM1.26: Red Bull^®^ Energy Drink + 1.26g of Calcium Citrate Malate; EDCaCM2.53: Red Bull^®^ Energy Drink + 2.53g of Calcium Citrate Malate; EDCaCM3.80: Red Bull^®^ Energy Drink + 3.80g of Calcium Citrate Malate; ED: Red Bull^®^ Energy Drink; DW: Deionized Water; OJCa: Orange juice with calcium Life Mix Baixa Caloria^®^.

### 3.2. Surface roughness

According to the Wilcoxon test, when comparing the surface roughness of the specimens before and after exposure to the drinks, only EDCaCM3.80 and DW groups did not present a significant difference (p > 0.05). The Kruskal Wallis test revealed significant difference (p < 0.001) between the drinks. Further analysis with the Mann-Whitney test allowed for identifying the specific groups where this significance occurred (p < 0.004). Among the modified drinks, EDCaPP1.43 exhibited less surface roughness than EDCaM1.66, and EDCaM1.66 showed greater roughness than EDCaCM1.26. Regarding the control groups, all modified drinks had less roughness than ED. Only EDCaPP1.43 did not present a significant difference compared to DW. When compared to OJCa, only EDCaM0.83 and EDCaM1.66 did not show a significant difference (Table 2).

**Table 2.** Inferential statistics of the roughness analysis of dental enamel specimens before and after exposure to drinks.

| Group | Mean BEFORE<br>(SD) | Mean AFTER<br>(SD) | p-value** |
| --- | --- | --- | --- |
| EDCaPP0.71 | 0,07<br>(0,01) | 0,09<br>(0,01) | 0,04 |
| EDCaPP1.43 | 0,05<br>(0,01) | 0,08<br>(0,02) | 0,04 |
| EDCaPP2.15 | 0,06<br>(0,01) | 0,09<br>(0,01) | 0,04 |
| EDCaM0.83 | 0,05<br>(0,01) | 0,10<br>(0,02) | 0,04 |
| EDCaM1.66 | 0,06<br>(0,02) | 0,11<br>(0,01) | 0,04 |
| EDCaM2.50 | 0,04<br>(0,01) | 0,09<br>(0,02) | 0,04 |
| EDCaCM1.26 | 0,05<br>(0,02) | 0,09<br>(0,01) | 0,04 |
| EDCaCM2.53 | 0,06<br>(0,02) | 0,09<br>(0,01) | 0,04 |
| EDCaCM3.80 | 0,06<br>(0,02) | 0,09<br>(0,02) | 0,08 |
| ED | 0,06<br>(0,01) | 0,29<br>(0,05) | 0,04 |
| DW | 0,05<br>(0,02) | 0,05<br>(0,02) | 0,69 |
| OJCa | 0,05<br>(0,02) | 0,16<br>(0,02) | 0,04 |
| p-value* | < 0,001 |  |  |

### 3.3. Surface microhardness

The ANOVA results showed significant effects for time (p < 0.0001), drink type (p < 0.0001), and their interaction (p = 0.0244). These findings indicate that both the evaluation time (initial and final) and the type of drink tested significantly influenced the microhardness values, with a significant interaction between the two factors (Table 3).

**Table 3.** Analysis of Variance (ANOVA) for the variable “microhardness” of dental enamel specimens.

| Source | DF | SS | MS | F | p-value |
| --- | --- | --- | --- | --- | --- |
| Time | 1 | 23037.0 | 23037.0 | 74.69 | < 0,0001 |
| Drink | 11 | 14938.7 | 1358.1 | 4.40 | < 0,0001 |
| Time x Drink | 11 | 7253.0 | 659.4 | 2.14 | 0.0244 |
| Error | 96 | 29608.5 | 308.4 |  |  |
| Total | 119 | 74837.3 |  |  |  |

DF: Degrees of Freedom; SS: Sun of Squares; MS: Mean Square

The Tukey HSD test revealed significant differences (p < 0.05) between the drink means (Table 4) showing that most of the modified drinks presented higher microhardness values compared to ED. This suggests that the modified drinks preserved more enamel structure, indicating less dental erosion than ED. Only EDCaM0.83, EDCaCM2.53 and EDCaPP0.71 did not show significant differences from ED. Furthermore, EDCaM2.50, EDCaPP2.15, EDCaM1.66 and EDCaPP1.43 also demonstrated better enamel preservation compared to the positive control, OJCa.

**Table 4.** Comparison of drink means using the Tukey HSD Test for the variable “microhardness” of dental enamel specimens.

| Group | Mean | Homogeneous groups |
| --- | --- | --- |
| DW | 358.17 | A |
| EDCaM2.50 | 352.97 | AB |
| EDCaPP2.15 | 342.86 | ABC |
| EDCaM1.66 | 337.77 | ABC |
| EDCaPP1.43 | 336.32 | ABC |
| OJCa | 334.01 | ABC |
| EDCaCM3.80 | 329.83 | BC |
| EDCaCM1.26 | 329.17 | BC |
| EDCaM0.83 | 325.98 | C |
| EDCaCM2.53 | 325.87 | C |
| ED | 322.65 | C |
| EDCaPP0.71 | 321.41 | C |

Different letters indicate significant differences between groups; DW: Deionized Water; EDCaM2.50: Red Bull® Energy Drink + 2.50g of Dicalcium Malate; EDCaPP2.15: Red Bull® Energy Drink + 2.15g of Calcium/Phosphorus/Potassium Complex; EDCaM1.66: Red Bull® Energy Drink + 1.66g of Dicalcium Malate; EDCaPP1.43: Red Bull® Energy Drink + 1.43g of Calcium/Phosphorus/Potassium Complex; OJCa: Orange juice with calcium Life Mix Baixa Caloria®; EDCaCM3.80: Red Bull® Energy Drink + 3.80g of Calcium Citrate Malate; EDCaCM1.26: Red Bull® Energy Drink + 1.26g of Calcium Citrate Malate; EDCaM0.83: Red Bull® Energy Drink + 0.83g of Dicalcium Malate; EDCaCM2.53: Red Bull® Energy Drink + 2.53g of Calcium Citrate Malate; ED: Red Bull® Energy Drink; EDCaPP0.71: Red Bull® Energy Drink + 0.71g of Calcium/Phosphorus/Potassium Complex

The interaction between time and drink (Table 5) further highlights the protective effect of most modified drinks on enamel structure, especially when compared to the final time of ED. EDCaM2.50, EDCaPP2.15, EDCaPP1.43, and EDCaM1.66 showed significantly higher microhardness values at the final time compared to ED (p < 0.05). When compared to OJCa, only EDCaPP0.71, EDCaM0.83, and EDCaCM2.53 did not show significant differences at the final time.

**Table 5.** Comparison of the means for the time x drink interaction using the Tukey HSD Test for the variable “microhardness” of dental enamel specimens.

| Time | Group | Mean | Homogeneous groups |
| --- | --- | --- | --- |
| Initial | EDCaM2.50 | 368.39 | A |
| Final | DW | 358.61 | AB |
| Initial | DW | 357.74 | AB |
| Initial | ED | 355.67 | ABC |
| Initial | OJCa | 354.99 | ABC |
| Initial | EDCaM1.66 | 352.24 | ABCD |
| Initial | EDCaPP2.15 | 351.15 | ABCD |
| Initial | EDCaCM3.80 | 344.92 | ABCD |
| Initial | EDCaPP1.43 | 344.40 | ABCD |
| Initial | EDCaCM1.26 | 344.09 | ABCD |
| Initial | EDCaCM2.53 | 340.51 | ABCD |
| Initial | EDCaM0.83 | 339.65 | ABCD |
| Final | EDCaM2.50 | 337.55 | ABCD |
| Final | EDCaPP2.15 | 334.56 | ABCD |
| Initial | EDCaPP0.71 | 329.54 | ABCDE |
| Final | EDCaPP1.43 | 328.24 | ABCDE |
| Final | EDCaM1.66 | 323.30 | BCDE |
| Final | EDCaCM3.80 | 314.74 | CDE |
| Final | EDCaCM1.26 | 314.26 | CDE |
| Final | EDCaPP0.71 | 313.28 | DE |
| Final | OJCa | 313.04 | DE |
| Final | EDCaM0.83 | 312.32 | DE |
| Final | EDCaCM2.53 | 311.24 | DE |
| Final | ED | 289.63 | E |

Different letters indicate significant differences between groups; EDCaM2.50: Red Bull® Energy Drink + 2.50g of Dicalcium Malate; DW: Deionized Water; ED: Red Bull® Energy Drink; OJCa: Orange juice with calcium Life Mix Baixa Caloria®; EDCaM1.66: Red Bull® Energy Drink + 1.66g of Dicalcium Malate; EDCaPP2.15: Red Bull® Energy Drink + 2.15g of Calcium/Phosphorus/Potassium Complex; EDCaCM3.80: Red Bull® Energy Drink + 3.80g of Calcium Citrate Malate; EDCaPP1.43: Red Bull® Energy Drink + 1.43g of Calcium/Phosphorus/Potassium Complex; EDCaCM1.26: Red Bull® Energy Drink + 1.26g of Calcium Citrate Malate; EDCaCM2.53: Red Bull® Energy Drink + 2.53g of Calcium Citrate Malate; EDCaM0.83: Red Bull® Energy Drink + 0.83g of Dicalcium Malate; EDCaPP0.71: Red Bull® Energy Drink + 0.71g of Calcium/Phosphorus/Potassium Complex

The %SMHL for enamel was calculated for all drinks (Fig. 3). When comparing the calcium formulations, the maximum difference in %SMHL between the formulations was 0.24%, 0.32% and 0.15%, respectively. Among the different calcium formulations, the calcium/phosphorus/potassium complex variation resulted in the lowest %SMHL. All specimens exposed to the modified drinks had lower %SMHL than those exposed to ED and OJCa.

**Fig 3.**
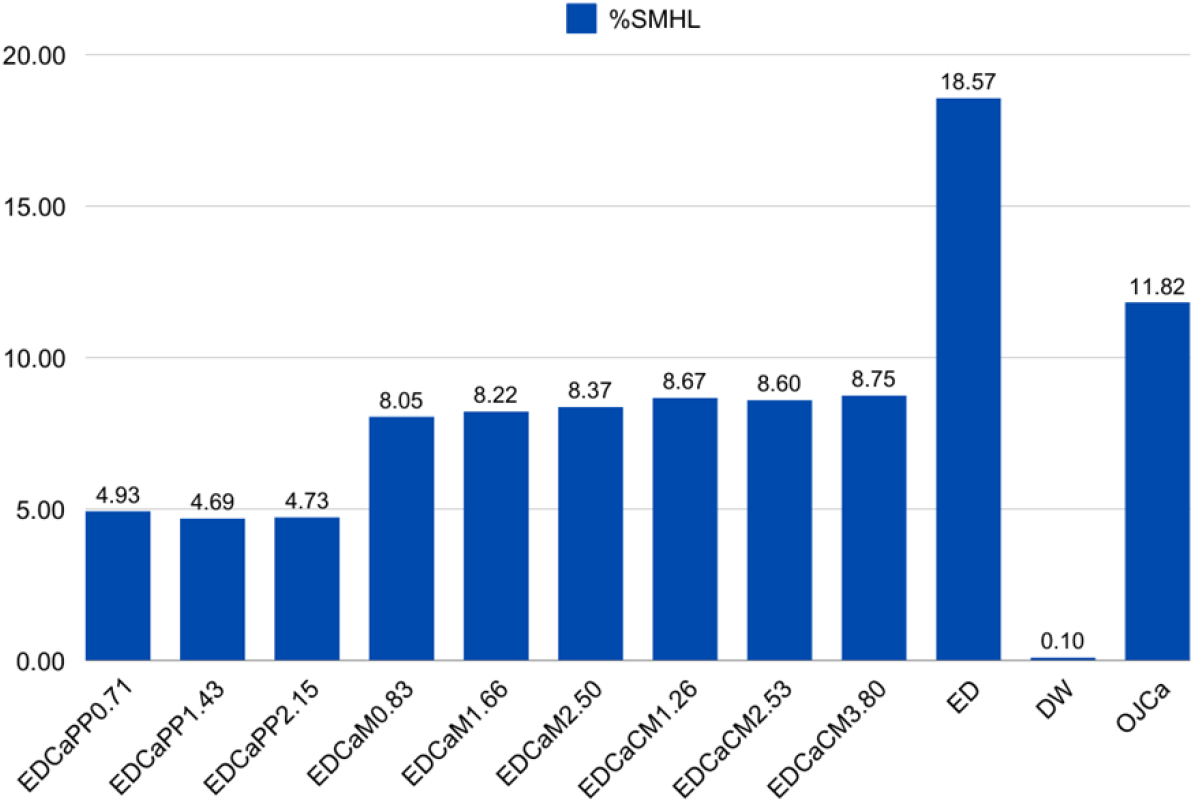
Column graph of percentage of surface microhardness loss (%SMHL) of enamel after exposure to drinks. EDCaPP0.71: Red Bull^®^ Energy Drink + 0.71g of Calcium/Phosphorus/Potassium Complex; EDCaPP1.43: Red Bull^®^ Energy Drink + 1.43g of Calcium/Phosphorus/Potassium Complex; EDCaPP2.15: Red Bull^®^ Energy Drink + 2.15g of Calcium/Phosphorus/Potassium Complex; EDCaM0.83: Red Bull^®^ Energy Drink + 0.83g of Dicalcium Malate; EDCaM1.66: Red Bull^®^ Energy Drink + 1.66g of Dicalcium Malate; EDCaM2.50: Red Bull^®^ Energy Drink + 2.50g of Dicalcium Malate; EDCaCM1.26: Red Bull^®^ Energy Drink + 1.26g of Calcium Citrate Malate; EDCaCM2.53: Red Bull^®^ Energy Drink + 2.53g of Calcium Citrate Malate; EDCaCM3.80: Red Bull^®^ Energy Drink + 3.80g of Calcium Citrate Malate; ED: Red Bull^®^ Energy Drink; DW: Deionized Water; OJCa: Orange juice with calcium Life Mix Baixa Caloria^®^.

## 4. Discussion

This study evaluated the erosive potential of an energy drink modified with different calcium formulations and concentrations on dental enamel. The results support that these modifications effectively reduce the erosive potential, leading to the rejection of the null hypothesis.

Among the factors influencing erosive potential, pH plays a key role. In this study, the addition of the calcium/phosphorus/potassium complex raised the drink’s pH, especially at higher concentrations, approaching the critical threshold for enamel dissolution (pH 5.5) [19], thus contributing to the reduced enamel erosion. In contrast, dicalcium malate and calcium citrate malate decreased the pH, likely due to the presence of malic and citric acids in these compounds, known for enhancing formulations acidity [20]. Beyond pH modulation, calcium in these formulations contributes to remineralization by restoring minerals lost from teeth through the enamel’s organic matrix [21]. This strategy has been widely adopted in the drink industry, particularly in plant-based drinks, to enhance nutritional value [22].

While prior studies have assessed the erosive effects of energy drinks, few have evaluated calcium supplementation within this specific drink category [23–25]. Our findings support these previous observations and highlight calcium as a promising additive to reduce dental erosion. Key parameters assessed in these studies also included drink pH, enamel roughness and microhardness [23–25].

Considering these crucial parameters, in the present study, drinks modified with calcium formulations resulted in significantly lower roughness values compared to ED and OJCa, indicating better enamel preservation. These results are consistent with prior evidence which suggests that the incorporating calcium into energy drinks can reduce surface roughness and overall enamel degradation [26].

Regarding microhardness analyses, specimens exposed to drinks with the highest concentration of dicalcium malate and the calcium/phosphorus/potassium complex presented significantly higher microhardness compared to those exposed to ED. Furthermore, the mean microhardness of specimens in these groups was also higher than that of OJCa. However, regarding %SMHL, all concentrations of the three tested calcium formulations demonstrated greater preservation compared to ED and OJCa, with calcium/phosphorus/potassium complex showing the best performance.

The protective effect observed is likely attributable to the high bioavailability of calcium, which plays a crucial role in reducing erosion caused by acidic drinks, including juices, carbonated drinks, and energy drinks [27]. This is further supported by the fact that other potential confounding factors were controlled in this study. While other strategies—such as adding green tea extract to energy drinks—have shown benefits in reducing teeth erosion by forming protective surface layers [28,29], such modifications have minimal impact on pH. In contrast, calcium not only protects against demineralization but also alters drink acidity, thereby providing dual benefit.

The calcium formulations tested in this study are viable and safe alternatives for drink industry [30–33]. In addition to their general health benefits and role in dental remineralization, they possess properties that further support their safety and effectiveness in drink modification [30–33]. These properties include high bioavailability, efficient absorption, and utilization by the body. Moreover, they do not alter sensory characteristics, shorten shelf life, or interfere with other nutrients in the formulation [30–33]. These factors are essential for achieving the nutritional objectives of the drink while ensuring consumer safety.

Despite the promising results demonstrated by the calcium formulations tested in this study, organoleptic tests were not performed to evaluate the sensory acceptability of modified drinks. These tests are essential for consumer-oriented development, as they assess flavor, aroma, color, and texture [34,35]. Additionally, enamel specimens were not exposed to saliva. While saliva plays a role in remineralization [28,29,36,37], its exclusion is justified in early-stage erosion models to isolate the effects of the tested variable [38].

Given these findings, future studies should focus on identifying the optimal calcium variation and the minimum effective concentration needed to reduce the erosive potential of energy drinks, particularly incorporating demineralization/remineralization cycles to better simulate real-life oral conditions. Moreover, *in vivo* studies are also necessary to evaluate the organoleptic properties and consumer acceptability of modified drinks. Enhancing the composition of these drinks could help protect oral health, especially as energy drink consumption continues to rise, along with its associated risk of dental erosion [39].

## 5. Conclusion

The addition of calcium formulations to the energy drink effectively reduced its erosive potential at all tested concentrations. Among them, the calcium/phosphorus/potassium complex showed the greatest protective effect, increasing pH and preserving enamel integrity.

## Data Availability

All relevant data are within the manuscript and its Supporting Information files.

## Acknowledgments

This research was supported by the Coordenação de Aperfeiçoamento de Pessoal de Nível Superior (CAPES) – Brazil and by the Programa de Apoio à Pós-Graduação (PROAP) – Brazil.

## References

[1] Silva ETC, Vasconcelos RG, Vasconcelos MG. Lesiones cervicales no cariosas: consideraciones etiológicas, clínicas y terapéuticas. Rev Cubana Estomatol 2019;56(4):1–15.

[2] Kanzow P, Wegehaupt FJ, Attin T, Wiegand A. Etiology and pathogenesis of dental erosion. Quintessence Int 2016;47(4):275–8.

[3] Huysmans MC, Chew HP, Ellwood RP. Clinical studies of dental erosion and erosive wear. Caries Res 2011;45:60–8.

[4] Huysmans MC, Young A, Ganss C. The role of fluoride in erosion therapy. Monogr Oral Sci 2014;25:230–43.

[5] Shellis RP, Addy M. The interactions between attrition, abrasion and erosion in tooth wear. Monogr Oral Sci 2014;25:32–45.

[6] Buzalaf MAR, Magalhães AC, Rios D. Prevention of erosive tooth wear: targeting nutritional and patient-related risks factors. Br Dent J 2018;224(5):371–8.

[7] Barac R, Gasic J, Trutic N, Sunaric S, Popovic J, Djekic P, et al. Erosive effect of different soft drinks on enamel surface in vitro: application of stylus profilometry. Med Princ Pract 2015;24(5):451–7.

[8] Carvalho TS, Schmid TM, Baumann T, Lussi A. Erosive effect of different dietary substances on deciduous and permanent teeth. Clin Oral Investig 2017;21(5):1519–26.

[9] Ogbeide UM, Irene DE, Okeri HA. Assessment of pH, titratable acidity, and caffeine content of some brands of energy drinks. J Sci Pract Pharm 2021;8:441–6.

[10] Martínez LM, Lietz LL, Tarín CC, García CB, Tormos JIA, Miralles EG. Analysis of the pH levels in energy and pre-workout beverages and frequency of consumption: a cross-sectional study. BMC Oral Health 2024;24:1–10.

[11] Carlo B, Barabanti N, Piccinelli G, Faus-Matoses V, Cerutti A. Microbiological characterization and effect of resin composites in cervical lesions. J Clin Exp Dent 2017;9:40–5.

[12] Inchingolo F, Dipalma G, Azzollini D, Trilli I, Carpentiere V, Hazballa D, et al. Advances in preventive and therapeutic approaches for dental erosion: a systematic review. Dent J 2023;11(274):1–20.

[13] Khan SA, Khalid N, Maqsood S, Tariq A, Hassan M. Association of carbonated drinks intake with dental erosion among dental students: a comparison between day scholars and hostel residents. Int J Contemp Med Res 2020;7(4):1–4.

[14] Chatzidimitriou K, Seremidi K, Kloukos D, Gizani S, Papaioannou W. The role of calcium in the prevention of erosive tooth wear: a systematic review and meta-analysis. Evid Based Dent 2024;25:55–55.

[15] Brazil. Ministério da Saúde. Agência Nacional de Vigilância Sanitária. Resolução RDC n. 719, de 01 de julho de 2022. Provides sanitary requirements for food preparation mixtures and ready-to-eat foods, https://www.in.gov.br/web/dou/-/resolucao-rdc-n-719-de-1-de-julho-de-2022-413364893; 2022 [accessed 2 May 2025].

[16] IOM / Institute of Medicine. Dietary reference intakes for calcium and vitamin D. Washington, DC: The National Academies Press; 2011.

[17] Vercelino CRMP, Costa ACS, Gurgel JA, Freitas KMS. Comparison of enamel surface roughness and color alteration after bracket debonding and polishing with 2 systems: a split-mouth clinical trial. Am J Orthod Dentofacial Orthop 2021;160(5):686–94.

[18] Al Kanhal H, Al Daij M, Al Kanhal N, Al Moither M, Albusair M, Baseer MA. Evaluation of the effect of different erosive drinks on teeth – Saudi Arabia, 2020. Med Sci 2021;25(116):2452–8.

[19] Delgado AJ, Ribeiro APD, Quesada A, Rodríguez LE, Hernández R, Wynkoop B, et al. Potential erosive effect of mouthrinses on enamel and dentin. Gen Dent 2018;66(3):75–9.

[20] Izquierdo-Llopart A, Saurina J. Characterization of sparkling wines according to polyphenolic profiles obtained by HPLC-UV/Vis and principal component analysis. Foods 2019;8(22):1–10.

[21] Roblegg E, Coughran A, Sirjani D. Saliva: an all-rounder of our body. Eur J Pharm Biopharm 2019;142:133–41.

[22] Rincon L, Braz ABR, Alencar ER. Development of novel plant-based milk based on chickpea and coconut. LWT 2020;128:1–29.

[23] Ostrowska A, Szymanski W, Kołodziejczyk Ł. Evaluation of the erosive potential of selected isotonic drinks: in vitro studies. Adv Clin Exp Med 2016;25(6):313–9.

[24] Damo DM, Arossi GA, Silva HA, Santos LH, Kappaun DR. Erosive potential of sports beverages on human enamel “in vitro”. Rev Bras Med Esp 2018;24(5):386–90.

[25] Silva JGVC, Martins JPG, Sousa EBG, Fernandes NLS, Meira IA, Sampaio FC, et al. Influence of energy drinks on enamel erosion: in vitro study using different assessment techniques. J Clin Exp Dent 2021;13(11):1076–82.

[26] Yildirim L. The effects of energy drinks on oral health and teeth. Braz J Implantol Health Sci 2023;5(5):4503–21.

[27] Jácome EVM, Bessa MSd, Borges BCD, Torres ACSP. Addition of substances to reduce the erosive potential of acidic beverages to tooth enamel: a scoping review. Int J Dent Hygiene 2024;22:758–68.

[28] Hamza B, Rojas SAP, Körner P, Attin T, Wegehaupt FJ. Green tea extract reduces the erosive dentine wear caused by energy drinks in vitro. Oral Health Prev Dent 2021;19:573–8.

[29] Blatter N, Hamza B, Attin T, Wegehaupt FJ. Supplementation of energy drinks with green tea extract: effect on in vitro abrasive/erosive dentin wear. Oral Health Prev Dent 2023;21:391–6.

[30] Andon MB, Peacock M, Kanerva RL, De Castro JA. Calcium absorption from apple and orange juice fortified with calcium citrate malate (CCM). J Am Coll Nutr 1996;15(3):313–6.

[31] Heaney RP, Dowell MS, Barger-Lux MJ. Absorption of calcium as the carbonate and citrate salts, with some observations on method. Osteoporos Int 1999;9:19–23.

[32] Chaturvedi P, Mukherjee R, McCorquodale M, Crowley D, Ashmead S, Guthrie N. Comparison of calcium absorption from various calcium-containing products in healthy human adults: a bioavailability study. FASEB J 2006;20:1063–4.

[33] Reinwald S, Weaver CM, Kester JJ. The health benefits of calcium citrate malate: a review of the supporting science. Adv Food Nutr Res 2008;54:219–346.

[34] Meilgaard MC, Civille GV, Carr BT. Sensory evaluation techniques. 4rd ed. Boca Raton: CRC Press; 2007.

[35] Dutcosky SD. Sensory analysis of foods. 5rd ed. Curitiba: PUCPRESS; 2019.

[36] Lussi A, Megert B, Shellis RP. The erosive effect of various drinks, foods, stimulants, medications and mouthwashes on human tooth enamel. Swiss Dent J SSO 2023;133(7/8):440–55.

[37] Aripin NFK, Zahid NI, Rahim MAA, Yaacob H, Haris PI, Rahim ZHA, et al. A review of salivary composition changes induced by fasting and its impact on health. Food Sci Hum Wellness 2024;13:50–64.

[38] Mylonas P, Austin RS, Moazzez R, Joiner A, Bartlett DW. In vitro evaluation of the early erosive lesion in polished and natural human enamel. Dent Mater 2018;34(9):1391–1400.

[39] Clapp O, Morgan MZ, Fairchild RM. The top five selling UK energy drinks: implications for dental and general health. Br Dent J 2019;226(7):493–7.

